# Leprosy stigma in the healthcare setting: The treatment-seeking journey of persons affected by Leprosy in Niger

**DOI:** 10.1101/2024.05.29.24308051

**Authors:** Amina P. Alio, Ibrahim Malam Mamane Sani, Yohanna Abdou, Moussa Gado, Issa Harouna, Bahadir Celiktemur

**Affiliations:** University of Rochester Medical Center, Rochester, New York, United States of America; The Leprosy Mission, Niamey, Niger; Université Abdou Moumouni, Niamey, Niger; Programme Nationale de Lutte contre la Lèpre, Ministère de la Santé, Niamey, Niger; IDEA-Niger, Maradi, Niger; The Leprosy Mission-Great Britain, London, United Kingdom

## Abstract

Leprosy-related stigma is deeply rooted in history and permeates every aspect of society, and stigmatization by healthcare professionals is no exception. This paper explores the experiences of persons affected by Leprosy with stigma in the healthcare setting throughout their treatment-seeking journey. We conducted one-on-one interviews with 112 individuals who experienced a Leprosy diagnosis and underwent treatment. Data was collected in July and August 2022, in 9 villages and 1 urban neighborhood in the region of Maradi, Niger. Over two-thirds of participants experienced some form of visible impairment due to Leprosy. Throughout their treatment-seeking journey, healthcare providers were reported to have stigmatized patients by using hurtful language, isolating patients, physically distancing from patients, or refusing to treat. The extent of the stigmatization depended on, first, whether Leprosy was recognized or not. Unfortunately, many healthcare providers did not detect the early signs of Leprosy, therefore, patients were then not able to obtain treatment prior to physical impairment. Second, the level of stigmatization varied based on the degree and type of visible physical impairments (e.g., missing fingers or toes, limbs, blindness). These negative experiences with healthcare workers made treatment-seeking and adherence to treatment regimen less likely for patients. Our findings confirm the presence of Leprosy stigma in the healthcare context in Niger. Increasing Leprosy knowledge and addressing stigma among healthcare professionals is key to early diagnosis and treatment of Leprosy.

## Introduction

Leprosy is a neglected tropical disease (NTD) that has been synonymous with disability and stigmatization throughout history. (1, 2) Caused by the bacillus Leprae, Leprosy attacks the nerves of the skin, causing visible alterations ranging from discoloration in spots, to boils. (3) Loss of sensation is the primary initial symptom distinguishing it from other skin conditions, making initial early diagnosis difficult without a blood test specifically for the bacillus. The disease develops slowly, with an average incubation period of three years. It can affect people of all ages and sexes, including children. Leprosy can be cured at any stage of the disease with multi-drug therapy (MDT) of antibiotics prior to it causing disability if detected and treated early. When left untreated, the consequences of Leprosy are lifelong and negatively affect the physical, social and emotional well-being of those affected and their families. The sequalae of Leprosy include serious impairment of the extremities, including hands, feet and the face. Because the loss of sensation is permanent, even after treatment the person affected by Leprosy is destined to a life of self-care to avoid further damage to desensitized skin. This self-care includes protection from the elements and injury. Because of the lack of sensation and not feeling any pain, wounds are often neglected until late stages that result in amputation of a limb. As a result, Leprosy has been associated with much stigma not only because of the beliefs associated with its transmission, but the physical impairment and disabilities that ensue.(3, 4)

Views about leprosy’s causal origins are two-fold. First, there is the fatalistic perception that the disease has both natural and divine origins, and that it is of a punitive nature for acts committed by the individual or a parent. The belief that Leprosy is contracted by one who is being punished or cursed by a divine power has been common in multiple religions and examples are recorded in their books (e.g., Bible, Torah, Kuran). (1, 4) In Niger, as in many African countries, Leprosy exists and people affected and their offspring are highly stigmatized and shunned from their communities of origin, forcing them to create new communities (often referred to as ‘colonies’) with other persons affected by the disease and their families. (5)

Leprosy is no longer considered a Public Health issue, but the disease persists in many developing countries like Niger. The Niger Ministry of Health reports that, out of about 4,000 individuals with skin conditions screened, 689 cases of Leprosy were treated, of which 259 were cured in 2022.(6) Leprosy remains a concern in Niger because new cases continue to be diagnosed (400 cases in 2023), even among children (10% of cases), indicating that the disease continues to be transmitted. We expect the real number of cases / hidden cases to be much higher. Additionally, the physical and social life-long impact on patients and their families render Leprosy a greater social issue in Niger.

Consequently, understanding the manifestations and drivers of Leprosy stigma, as described by the World Health Organization (WHO) stigma framework, (7) is key to informing interventions to reduce stigma and improve the lives of those affected and their communities. In the case of Leprosy, early detection and adherence to treatment is key to preventing physical impairment (e.g., missing fingers, toes, limbs, disfigurement, blindness) and associated disability (experiences with environmental and social barriers/structures).(8) Stigma is a concept embodying elements of labeling, stereotyping, ‘othering’, separating, status loss and discrimination occurring within a situation of power. (9) Stigmatization by family and community members, as well as by healthcare providers is the primary barrier to patients accessing screening services and treatment. Studies on community stigma in African countries are multiple, however, very few have focused on the problem of stigma by healthcare providers. The few studies that describe stigmatization by healthcare providers highlight the need to better understand the phenomenon and to address the problem. (10-12) Mirroring their society, healthcare providers have been known to discriminate against patients with Leprosy or those living with the sequelae. A study in Ghana recognized that healthcare providers were afraid of Leprosy and therefore avoided patients who had the disease because of the visible sequelae. (13) A study in Nepal, although it does not directly address discrimination by healthcare providers, described the effects of stigma on healthcare seeking behaviors. (14) Stigma was found to affect where patients sought treatment, with many of them opting to obtain treatment from spiritual or traditional healers, which delays their treatment and increases chances of resulting impairments and disability.

### Objectives

Despite advancements in knowledge about the disease and its treatment, leprosy stigma persists in society, and particularly in healthcare settings. Misconceptions about the disease and its transmission still abound, leading to discrimination and prejudice against leprosy patients and patients living with the sequelae of the disease needing treatment for co-morbidities. Given the gap in the literature about the topic and the context of Niger, this paper presents results of a study exploring the experiences of persons affected by Leprosy with stigma throughout their treatment seeking journey, highlighting stigmatization in the healthcare context.

## Methods

This is an exploratory, qualitative study using a descriptive, narrative approach (15) to understanding the experiences of persons infected with Leprosy throughout their treatment-seeking journey. Our specific objectives were to understand the experiences of patients with Leprosy, from diagnosis to treatment and cure, and to identify the manifestations and drivers of stigma experienced in the healthcare setting.

Participants were recruited via the Maradi regional chapter of IDEA (Intégration, Dignité, Economie en Avant), an international organization that brings together persons and communities affected by disability, including those affected by Leprosy. IDEA is focused on the social integration and economic advancement of persons affected by disability and has chapters across countries globally. IDEA Niger was instrumental as a community partner and stakeholder in this study.

In July and August 20022, we conducted individual interviews with 112 participants in 10 communities (9 villages and one urban neighborhood) of persons affected by Leprosy and their children for multiple generations. Interviews solicited individual narratives of their experiences with stigma throughout their journey to treatment and healing.

IDEA Maradi discussed the study with community leaders from all 10 communities, after which announcements were made at community gatherings, via telephone messaging, word of mouth and WhatsApp. Participants meeting the criteria of having been diagnosed with Leprosy at any point in their lives and who were 18 years or older, were invited to come to a specific community center or square at a specific date and time. Interviews were audio recorded with the informed consent of the participants. The interview guide was semi-structured in order to maintain consistency between the 5 trained interviewers. Structured questions included general demographic characteristics, information about the sequelae of Leprosy (i.e., impairments and disabilities), and open-ended questions were about their experiences beginning with the diagnosis of Leprosy to their current status. Probes focused on manifestation of stigma in the healthcare setting and the drivers of stigma. Interviews lasted on average 45 minutes (range of 35 to 70 minutes). Interviews were conducted in Hausa, the primary language of the region, and known to all the interviewers.

Interview audios were transcribed by the interviewers using direct translation into French. Transcripts were verified for accuracy by two of the researchers who met with the project advisory team of stakeholders to discuss the accurate translation of certain local concepts. Transcripts were uploaded into Dedoose and coded using a descriptive, narrative analysis approach (15) within the context of the phenomenon of interest, the treatment-seeking stories of persons affected by Leprosy. The narrative focused on describing common experiences in the journey to healing. Multiple coders were used, and disagreements discussed until consensus was reached. Members of IDEA who themselves are affected by Leprosy participated in discussions to validate or correct researchers’ understanding of the data and its interpretation.

### Ethical Considerations

The study was approved by the Ministère national de l’éducation supérieure et de la recherche in Niamey, Niger, and by the University of Rochester Research Subject Review Board, New York, USA.

## Results

Of the 112 persons affected by Leprosy interviewed, 47 were female (42%). Though most were not sure of their exact age, participants were older adults, estimated to be between 35 to over 70. Most (64%) were diagnosed late or where not initially adherent to treatment and had visible sequelae of the disease, including disfigurement to the feet (n=22), hands (n=33), face (n=15), both hands and feet (n=2), and other impairments (n=17) like the loss of a limb, for example. Almost all had no formal schooling (95%) but 30% of them had attended literacy classes as part of community activities by NGOs and CBOs. Most participants were married (71%) or widows/widowers (20%). They had an average of 4.7 children each, ranging from no children to 19. Participants made ends meet primarily by farming, breeding animals, small commerce/business, or other manual labor. Over a third (36%) reported begging as their main source of income (Table 1).

**Table 1.**
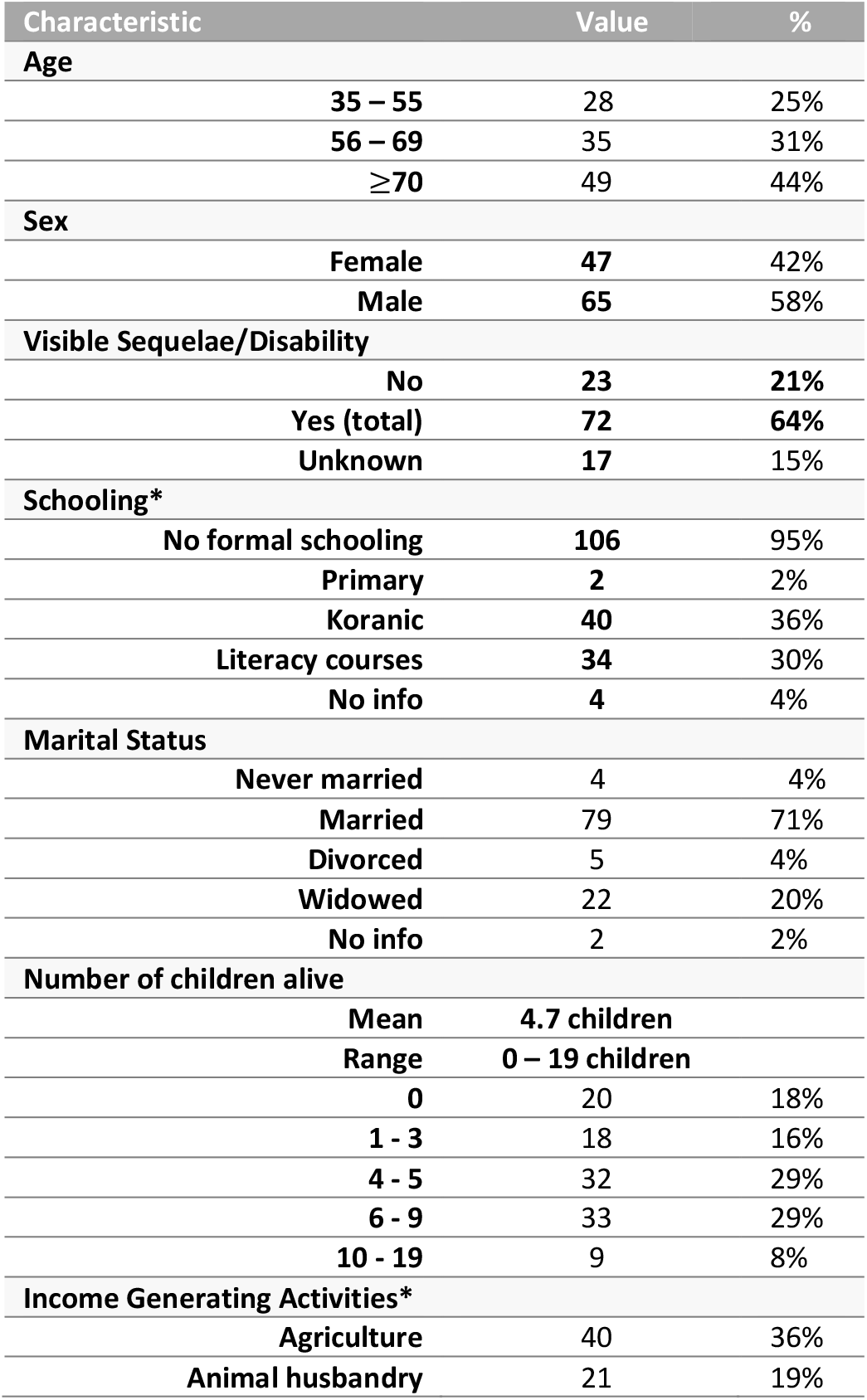
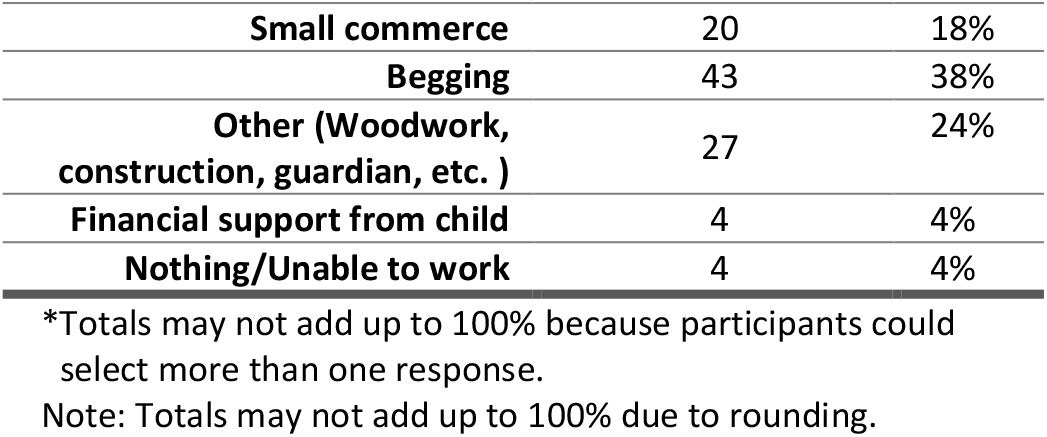
Participant characteristics (N= 112)

### The beginning of the journey: Diagnosis of Leprosy

The narratives followed 2 primary paths: those who were diagnosed and treated in the early stages of the disease, and those who were not and therefore experienced physical impairment and/or disabilities. Most noticed initial symptoms like spots on their skin and immediately sought care at a clinic or from a traditional healer. Unfortunately, for the majority, Leprosy was not immediately diagnosed and they were treated for other skin conditions. When the symptoms became worse, they sought care from traditional healers to no avail, and then turned to formal medical clinics or hospitals. Because the symptoms had become worse, Leprosy became more recognizable and was finally diagnosed. For most, they were diagnosed with Leprosy as adolescents or young adults, although the initial symptoms indicate they most likely had had it for a time.

For about a third of the participants, the diagnosis occurred early as a result of someone in their social circle recognizing the symptoms because they had seen it within themselves or someone they knew. A male participant who was diagnosed as a child remembers: *“I was visiting my uncle and he told me that I needed to go to this special hospital to be treated for my skin condition because it could be Leprosy. So they took me to [Likita] Hospital*.*”* The hospital mentioned [given the pseudonym of Likita] is one of the largest hospitals with a specialized Leprosy treatment center in the country. Those who were diagnosed early or eventually all ended up at Likita where they were diagnosed with Leprosy and treated.

The reactions to the diagnosis were similar across participants. They felt it was “*worse than a death sentence*,” because they would be living with a disease that would change the course of their lives. All ended up leaving their home and community of origin, for various reasons, the primary themes being:

- rejection by their family or community: “*When my parents found out they were afraid and sent me away. But everywhere I went people treated me badly until someone told me about Likita*.”
- fear of rejection by their family and community: “*I knew if people find out what I have they will be afraid of me or think something bad about me*.”
- fear of the social consequences on their family: “*I was married and didn’t want my husband and children to suffer because of me, so I left my husband and [young] child. I left my home to come to [Likita] where I started a new life*.”
- the desire to find treatment at the Likita Hospital: *“My parents took me to Likita because they knew someone who was treated there. They left me with them and returned home*.”

### Treatment Seeking

All participants described doing all they could to treat their newly diagnosed condition of Leprosy. To them and their parents (for those who were children), that meant seeking both spiritual/traditional and modern medicine. Because Leprosy is seen as having an important spiritual aspect, medical treatment for the physical symptoms was not considered sufficient: the perception was that all aspects of the disease needed to be addressed for healing. Once diagnosed with Leprosy, going to the spiritual, traditional healer was purposeful and they finally had a specific reason for seeking healing. This was done parallel to the treatment at Likita for most. A few – mostly those who were younger at the time - quickly abandoned the harsh traditional treatment that consisted of incantations and very bitter potions to drink, and specially made ointments for the affected areas of the skin. One man describes his experience and as a child: *“It was so bitter! I didn’t want to drink it anymore. It made my stomach sick, and I would vomit. So I would cry and refuse even when people would tell me I needed it*.*”* Of note is that participants described positive experiences with the healers themselves, if not the treatment. The traditional healers were not stigmatizing of the patients that came to them with a diagnosis of Leprosy. They did not fear them or reject them, as it seemed to have been a spiritual matter that they were equipped and ready to deal with.

Parallel to the traditional treatment, many also sought medical treatment. Unfortunately for over 60% of our sample, the treatment came after the visible physical impairments and disabilities. The few who sought treatment in local health centers were put on treatment or referred to the few regional hospitals with medication for Leprosy. However, their experiences were not as positive as with the traditional healers. Some talked about clinicians refusing to treat them in a manner that was full of disgust or pity for their plight. The feelings of pity were, as the patients perceived it, because they were suffering the divine or spiritual wrath for something they of their family did. *“They sent me to the hospital in Tahoua and felt sorry for my condition. It is God’s doing*.*”* They perceived this attitude of pity as hurtful and bringing up feelings of shame for having Leprosy. Others described their interactions with healthcare workers as very negative, including being isolated from other patients, or the workers keeping a distance from them for fear of contagion. “*They didn’t want to touch me. They were afraid to catch what I had*.” These reactions of pity, fear and disgust were common among the study participants’ experience with the healthcare workers of all levels, from the clinicians to the staff. The health centers that treated them made it difficult to return regularly for their medication. *“They gave me pills for 2 months and told me to come back but I didn’t. I continued with my traditional treatment after the pills finished*.*”* When asked why he didn’t return, the man explained “*I just didn’t. I didn’t want to feel ashamed*.*”*

Eventually, all the patients ended up at the Likita referral hospital for Leprosy. The hospital, located in the far south-east of the country, includes a primary focus on the treatment of persons with Leprosy. The patients were either referred by someone they knew who recognized the signs of Leprosy, or by a clinician who diagnosed them or suspected a Leprosy diagnosis but was unable to confirm. The patients described the treatment they received in Likita Hospital as holistic, dealing with the physical, emotional, social, and spiritual aspects. “*They gave me medication and allowed me to stay there until my treatment was finished. I was there for many months*.” “*At Likita I found many others like me. Some had just arrived; some had been there for a while*.*” “They explained to me my condition and how to take care of my feet*.*” “They helped me with my emotions. We could talk to them or to other people like me anytime*.*” “I was feeling bad, but everybody was so nice and accepting of me*.*”* At Likita, they found a community of people who shared similar experiences and staff who cared for their well-being beyond Leprosy. “*The [Likita staff] really took their time and told me I could stay there until I was healed. So, my family left me there to go back to our home region. I’m still here after many years. I married and had children and I’m still here in this community*.*”*

As they live with the sequelae of Leprosy, participants with visible impairments and disabilities describe avoiding health centers outside of Likita to evade being stigmatized and discriminated against by healthcare workers. Though cured of the disease, they must suffer the long-term social effects of Leprosy, including fear, disgust and pity. *“We only go to Likita when we’re sick with anything, or when I have a sore that won’t heal*.*” “Likita is our hospital. Even if I go to the clinic here, when they see we [have leprosy] they automatically tell us to go to Likita. Even if we finished our Leprosy treatment*.*”*

Meeting a community of people sharing the same illness and social fate, the patients intermarried, created families and settled in the region in multiple settlements that are now villages and urban neighborhoods. These communities of persons affected by Leprosy, including patients, former patients and their offspring have become a safe haven for the newly infected who come from the various regions of Niger and northern Nigeria for treatment.

## Discussion

Leprosy stigma in healthcare settings remains a significant issue despite advancements in medical understanding and treatment options. Historically, leprosy, also known as Hansen’s disease, has been associated with fear, misunderstanding, and social exclusion. (16) While medical knowledge has advanced, the stigma surrounding leprosy continues to persist, including within healthcare settings.

One of the primary reasons for leprosy stigma in healthcare settings is the lack of education and awareness among healthcare professionals. (14, 17) Due to its historical associations with impairment and contagion, healthcare providers in our study harbored misconceptions about leprosy, leading to discriminatory attitudes and behaviors towards patients with the disease. In our study, this manifested as reluctance to provide care, segregation of leprosy patients from other patients, or even refusal to touch or examine them. Although little research has been published on discrimination in healthcare setting, much has been established regarding societal stigma based on misconceptions about Leprosy. (10, 16)

In particular, the physical manifestations of leprosy, such as skin lesions and impairments, evoked fear and/or revulsion among healthcare workers according to our findings, further exacerbating stigma. This fear led to avoidance of close contact with leprosy patients or neglect of their medical needs. Furthermore, the stigma surrounding leprosy in healthcare settings can have detrimental effects on patients’ mental health and well-being. Being subjected to discrimination and prejudice from healthcare providers led to feelings of shame, embarrassment, and low self-worth among leprosy patients. This, in turn, deterred them from seeking timely medical care or adhering to treatment regimens, resulting in permanent damage to their skin, face, hands and feet, resulting from delayed diagnosis, delayed treatment and non-adherence to their treatment regimen.

Alongside stigma towards leprosy and those with the disease, our findings highlight the important facet of lack of knowledge of leprosy etiology among healthcare providers. Indeed, the majority of participants who first went to clinics or hospitals outside of the main regional hospitals were not immediately diagnosed when the symptoms were limited to the skin, when disability could have been prevented. About a third were lucky enough to have someone with personal experience of the disease to refer them to Likita for correct diagnosis and treatment. This highlights the importance of training clinicians in how to recognize the early signs of Leprosy and how to treat it, given the medication is available in Niger. This has been recognized in the country and is one of the goals of the national Leprosy control program.(6) Early diagnosis and treatment would be lifechanging for persons affect by Leprosy, as they would not suffer from the physical consequences, and the stigma towards them would be greatly diminished. The participants who had no visible sequelae were better able to infiltrate society outside of their communities, although they lived with the fear of being found out. Should it be known that they had had Leprosy, there would be reticence to have them integrated fully and they would be stigmatized in various ways, ranging from mockery and insulting name calling to refusal to give their daughter’s hand in marriage.

In addition to healthcare providers, stigma within healthcare settings can also be perpetuated by institutional policies and practices. For example, having a large hospital like Likita for the care of persons with Leprosy, while being a place for specialized care may have the unintended consequence of reinforcing the idea of segregation and otherness. This could be seen in our findings: the patients found safety in perpetuating the idea that Likita is their hospital, as is the only place they feel welcomed. For these patients “othered,” Likita has become a safe place for healing and overall well-being, showing adaptation to the lifelong and intergenerational stigmatization of Leprosy. At the same time, Likita has become a stigmatized place by the larger society, with anyone being from Likita is treated as a person affected by Leprosy.

Addressing leprosy stigma in healthcare settings requires a multi-faceted approach that includes increasing healthcare provider knowledge of Leprosy, addressing Leprosy stigma among healthcare workers, and ensuring institutional policies emphasize the rights to care of patients. (11, 18) First and foremost, healthcare providers need to be educated about leprosy, its causes, transmission, and treatment. Training programs should focus on dispelling myths and misconceptions about the disease and promoting empathy and compassion towards patients. Additionally, healthcare facilities should implement policies that promote inclusivity and non-discrimination, such as integrating leprosy care into general healthcare services and ensuring equal access to treatment for all patients at all stages of the disease.

Training of healthcare provider should also include how to address stigma at the different points of the treatment-seeking journey of patients. For example, when diagnosing Leprosy, healthcare workers can be trained to provide counseling (similar to HIV diagnosis)(19) to address the fear of the disease and its social consequences while emphasizing the importance of adherence to treatment regimen for preventing impairment. For those diagnosed late, healthcare workers can be sure to address any social/environmental factors causing disability as a result of their impairment, while emphasizing the importance of adherence to treatment regimen and self-care needed to prevent further physical damage. At any point in the patient’s treatment process, most important is for them to model acceptance and compassion.

### Limitations

This study is limited in that it only represents the views of patients about their experiences in healthcare centers, as was the goal of this study. We do not represent the perspective of healthcare professionals. Future research may focus on measuring stigma among healthcare professionals to better understand their views about the disease and the people affected. Nevertheless, understanding the experiences of patients is most important to identify the existence of stigma and call for interventions to address stigma in the healthcare context, especially since it affects patient treatment-seeking behaviors.

## Conclusions

Leprosy stigma in healthcare settings persists due to a combination of lack of knowledge and fear. Addressing this stigma requires concerted efforts to educate healthcare providers, reform institutional practices, and engage communities in promoting acceptance and inclusion. Traditional/spiritual healers should be considered when planning interventions for Leprosy screening, as they can play an important role in helping to identify Leprosy in its early stages. Training of healthcare providers is also key to helping combat stigma and ensuring that Leprosy is diagnosed and treated early, sparing patients lifelong consequences of impairments and resulting disabilities. As stigma among healthcare providers stems partly from fear of contagion, increase in knowledge about the fact that Leprosy is one of the least contagious infectious diseases would help alleviate these fears. The example of the Likita Hospital staff’s treatment of Leprosy patients is evidence that trained healthcare workers and an approach to care that includes addressing the social consequences of Leprosy can lead to better detection, treatment, and the overall well-being of patients. By challenging stigma and fostering empathy and understanding among healthcare providers, we can ensure that all patients, including those affected by leprosy, receive equitable healthcare and support.

## Data Availability

Data are available upon reasonable request to the authors.

## Acknowledgement

Many thanks to the individuals who so willingly shared their stories.

